# Is artificial intelligence for medical professionals serving the patients? Protocol for a mixed method systematic review on patient-relevant benefits and harms of algorithmic decision-making

**DOI:** 10.1101/2024.03.27.24304965

**Authors:** Christoph Wilhelm, Anke Steckelberg, Felix G. Rebitschek

## Abstract

**Background:** Algorithmic decision making (ADM) utilizes algorithms to collect and process data and develop models to make or support decisions. Advances in artificial intelligence (AI) have led to the development of support systems that can be superior to medical professionals without AI support in certain tasks. However, whether patients can benefit from this remains unclear. The aim of this systematic review is to assess the current evidence on patient-relevant benefits and harms when healthcare professionals use ADM systems (developed using or working with AI) compared to healthcare professionals without AI-related ADM (standard care) - regardless of the clinical issues. Furthermore, for interpreting collected evidence and analysing preconditions for the implementation of AI-related ADM in healthcare, experts from research, practice, and regulation will be interviewed.

**Methods:** Following the PRISMA statement and the MECIR standards for reporting systematic reviews, MEDLINE and PubMed (via PubMed), EMBASE (via Elsevier), IEEE Xplore, CENTRAL will be searched using English free text terms in title/abstract, Medical Subject Headings (MeSH) terms and Embase Subject Headings (Emtree) fields. Additional studies will be identified by contacting authors of included studies and through reference lists of included studies. Grey literature searches will be conducted in Google Scholar. Risk of bias will be assessed by using Cochrane’s RoB 2 for randomised trials and ROBINS-I for non-randomised trials. Transparent reporting of the included studies will be assessed using the CONSORT-AI extension statement. Following the SRQR statement, semi-structured interviews will be conducted and analysed with the help of a qualitative content analysis according to Mayring. Based on the research questions and the findings of the systematic review, the study and interview guide will be developed a priori.

**Discussion:** It is expected that there will be a substantial shortage of suitable studies that compare healthcare professionals with and without ADM systems concerning patient-relevant endpoints. This can be attributed to the prioritization of technical quality criteria and, in some cases, clinical parameters over patient-relevant endpoints in the development of study designs. Furthermore, it is anticipated that a significant portion of the identified studies will exhibit relatively poor methodological quality and provide only limited generalizable results.

**Systematic review registration:** This study is registered within Prospero (CRD42023412156).

## Background

Artificial Intelligence (AI) is a broad term referring to the field of computer science that develops algorithms mimicking human cognitive functions such as learning, perception, problem-solving, and decision-making (McCarthy et al. 2006: 5-7). AI encompasses various approaches, including rule-based systems, machine learning (ML), deep learning, and many others. It comprises a range of technologies and techniques, including algorithmic decision-making (ADM) (Graili et al. 2021: 1). ADM refers to the process of using algorithms to gather, process and model input data to inform or make automated decisions. Feedback from these decisions can then be used by the system to improve itself (Araujo et al. 2020: 612). An ADM can take various forms depending on how it is framed and presented to the user or decision subject. It can be a simple algorithm that has been known and used for decades, such as classification trees (von Winterfeldt & Edwards 1986), or a more complex system like a recommender or AI that can provide recommendations to human decision-makers, nudge its users in a certain direction, or perform fully automated decision-making processes without human involvement (Araujo et al. 2020: 613). To sum up, artificial intelligent related algorithmic decision-making systems (AI-related ADM) are decision support systems that either apply AI (e.g., relying on ML models) or have been developed with the help of AI.

Recent advances in AI have resulted in the development of more complex and sophisticated systems that can outperform humans in certain tasks. One such example is AlphaZero, a deep learning algorithm developed by DeepMind, which uses reinforcement learning to learn how to play games like chess, shogi, and Go without being explicitly programmed. In fact, AlphaZero has convincingly beaten both human world champions and world champion computer applications in these games, simply by inputting the rules of the game (Silver et al. 2018: 3-4). Another notable AI system is ChatGPT, developed by OpenAI, which is a prototype text-based dialogue system that can generate human-like text and imitate writing essays and business plans (Hughes 2023). It can also analyse and write code in various programming languages, making it a valuable tool for debugging and code improvement (OpenAI 2023). Recently, ChatGPT was evaluated for its clinical reasoning ability by testing its performance on questions from the United States Medical Licensing Examination, where it scored at or near the passing threshold on all three exams without any special training or reinforcement (Kung et al. 2023).

These advances in AI seem to have enormous potential to transform many different fields and industries, which begs the question: will AI do so in healthcare?

In clinical trials, AI systems have already shown potential to help clinicians make better diagnoses (Bahl et al. 2018, Li et al. 2021), help personalise medicine and monitor patient care (Jiang et al. 2017, Ciervo et al. 2019), and contribute to drug development (Ekins et al. 2019). However, successful application in practice is limited (Panch et al. 2019: 77) and potential issues that may be responsible for this gap between research and practice should be revealed by our work.

By searching PubMed for the term “artificial intelligence”, we found over 2,000 systematic reviews and meta-analyses published in the last 10 years, with a yearly increasing trend. These include several reviews conducted in the area of AI in healthcare that provide an overview of the current state of AI technologies in specific clinical areas, including AI systems for breast cancer diagnosis in screening programmes (Freeman et al. 2021), ovarian cancer (Xu et al. 2022), early detection of skin cancer (Jones et al. 2022), COVID-19 and other pneumonia (Jia et al. 2022), prediction of preterm birth (Akazawa & Hashimoto 2022), or diabetes management (Kamel et al. 2022). Other reviews have focused on comparing clinicians and AI systems in terms of their performance to show their capabilities in a clinical setting (Shen et al. 2019, Nagendran et al. 2020, Liu et al. 2019).

Although these reviews are crucial to the further development of AI systems, they offer little insight into whether patients actually benefit from their use by medical professionals. Indeed, these studies focus on the analytical performance of these systems, rather than on healthcare-related metrics. In most of the studies mentioned here, the underlying algorithms have been evaluated using a variety of parameters, such as the F1 score for error classification, balanced accuracy, false positive rate, and area under the receiver operating characteristic curve (AUROC). However, measures of a system’s accuracy often provide non-replicable results (McDermot et al. 2021: 4), do not necessarily indicate clinical efficiency (Keane & Topol 2018: 1), AUROC does not necessarily indicate clinical applicability (Halligan et al. 2015: 935) and in fact, none of these measures reflect beneficial change in patient care (Brocklehurst et al. 2017: 1727, Shah et al. 2019: 1).

To summarise, as with any other new technology introduced into healthcare, the clinical effectiveness and safety of AI compared to the standard of care must be evaluated through properly designed studies to ensure patient safety and maximise benefits while minimising any unintended harm (Park et al. 2020: 328). Therefore, **a critical analysis of patient-relevant outcomes is needed, especially the benefits and harms of decisions informed by or made by AI systems**.

To this end, this review goes beyond previous studies in several ways. First, we study clinical AI systems that enable algorithmic decision making (AI-related ADM) in general and therefore do not limit ourselves to selected clinical problems. In particular, we focus on machine learning systems that infer rules from observations. Although we omit rule-based systems, we apply the term AI throughout our work because it is often incorrectly and redundantly used for ML and deep learning in the literature we study. Second, we focus on studies that report patient-relevant outcomes that, according to German Institute for Quality and Efficiency in Healthcare (IQWiG 2022: 44) describe, how patients feel, how they can perform their functions and activities or if they survive. These may include, for example, mortality, morbidity (with regard to complaints and complications), length of hospital stay, readmission, time to intervention and health-related quality of life. Third, we focus only on studies that compare medical professionals supported by AI-related ADM systems with medical professionals without AI-related ADM systems (standard care). By doing so, this review provides an overview of the current literature on clinical AI-related ADM systems, summarises the empirical evidence on their benefits and harms for patients and highlights research gaps that need to be addressed in future studies.

If clinical AI systems are found to have a well-balanced benefit-to-harm ratio, their implementation into practice can be considered. However, effectively deploying these systems presents a separate frontier. Specific challenges that need to be addressed include, data privacy and security, usability and lack of algorithm transparency (e.g., in the case of proprietary systems), regulations and policies, the changing nature of healthcare work, the risk of harm from system errors, and at the individual level, physician and patient awareness, education, and trust in these systems (Choudhury et al. 2020: 3, He et al. 2019: 33, Yu et al. 2018: 720).

To outline these challenges and framework conditions, particularly in the German healthcare system, and to consider the empirical data collected in this review, **it is necessary to consult experts from various disciplines to use their experiences, knowledge, and perceptions of the benefits and risks of implementing AI-related ADM systems**. Therefore, we will bring together experts from institutions responsible for auditing the safety of AI systems, audit bodies that support implementation processes under scientific and regulatory conditions in the healthcare sector, and bodies that combine practical healthcare work and research with AI systems.

## Objectives

The aim of this review is to systematically assess the current evidence on patient-relevant benefits and harms of ADM systems which are developed or used with AI (AI-related ADM) to support medical professionals compared to medical professionals without this support (standard care) (Study 1). Furthermore, we will elicit expert assessments of our findings and the benefits and risks of the implementation of AI-related ADM in healthcare, which they perceive (Study 2).

### Study 1, Systematic review

1. Are there studies that compare patient-relevant effectiveness of AI-related ADM for medical professionals compared to medical professionals without AI-related ADM?
2. Do these studies show adequate methodological quality and are their findings generalisable?
3. Can AI-related ADM systems help medical professionals to make better decisions in terms of benefits and harms for patients?

### Study 2, expert interviews

1. How do experts assess the overall benefit-to-harm ratio of identified AI-related ADM systems?
2. In which medical area of competences (diagnosis, treatment, prognosis, prevention) would patients in Western healthcare systems particularly benefit from AI-related ADM systems?
3. What are the consequences (e.g., side effects) of implementing AI-related ADM systems in healthcare?
4. What are the main challenges that hinder the implementation of AI-related ADM systems in clinical practice and what improvements could be suggested?
5. What are the regulatory requirements for implementation of AI-related ADM systems in clinical practice?

## Methods/Design

In accordance with the Preferred Reporting Items for Systematic Review and Meta-Analysis Protocols (PRISMA-P) statement (Moher et al. 2015), the study protocol for this systematic is registered on the International prospective register of systematic reviews (PROSPERO) database (CRD42023412156). If necessary, post-registration changes to the protocol, will be detailed under the PROSPERO record with an accompanying rationale.

To answer the research questions presented, a systematic review in mixed-method design will be conducted, which will be presented into two parts.

### Study 1: Systematic review

We will follow the Preferred Reporting Items for Systematic Reviews and Meta-Analyses (PRISMA) statement (Page et al. 2021) and the Methodological Expectations of Cochrane Intervention Reviews (MECIR) standards (Higgins et al. 2021).

### Searches

We will search systematically using English free text terms in title/abstract, Medical Subject Headings (MeSH) terms and Embase Subject Headings (Emtree) fields for various forms of keywords related to ‘artificial intelligence’ and relevant subcategories of computer generated and processed decision-making algorithms, ‘medical professionals’ and keywords describing effectiveness parameters and outcomes as well as preferred study types. Based on the block building approach, keywords and terms are combined using the Boolean operators AND and OR and progressively checked for relevant hits.

### Databases to be used for searches

MEDLINE and PubMed (via PubMed), EMBASE (via Elsevier), Institute of Electrical and Electronics Engineers (IEEE) Xplore, will be searched for peer-reviewed articles as well as ClinicalTrials.gov and ICTRP (via CENTRAL) for ongoing trials and protocols.

To reduce potential publication bias, additional studies will be identified by contacting authors of included studies, contacting experts in the field, and through reference lists of relevant studies. Grey literature searches will be conducted in Google Scholar. For this purpose, the keywords used in the systematic search will be used in different combinations, as well as their German equivalents. Google Scholar will be searched up to the 10th hit page. The detailed search strategy for each database will be reported under the PROSPERO record once the searches have been conducted.

### Search strategy

We developed our search strategy using the PICOS scheme (Table 1).

**Table 1:**
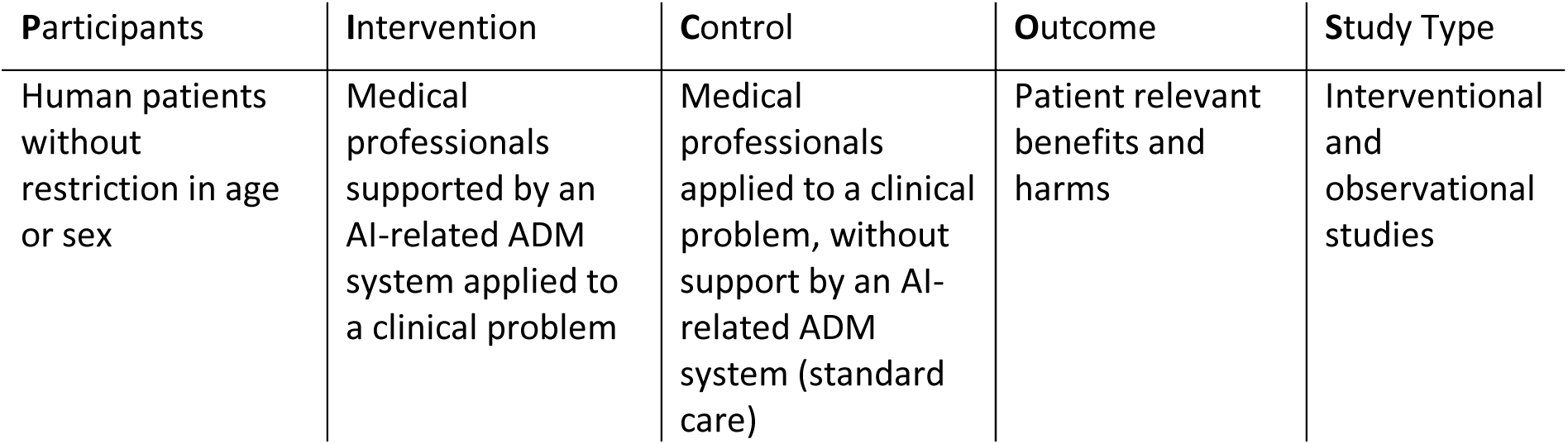
PICOS scheme.

| Participants | Intervention | Control | Outcome | Study Type |
| --- | --- | --- | --- | --- |
| Human patients without restriction in age or sex | Medical professionals supported by an AI-related ADM system applied to a clinical problem | Medical professionals applied to a clinical problem, without support by an AI-related ADM system (standard care) | Patient relevant benefits and harms | Interventional and observational studies |

While doing preliminary searches for basic literature in Medline and PubMed (via PubMed), we noticed that study conductors from different scientific fields (e.g., computer scientists) used different terms for the intervention outcomes we were looking for. In addition, some studies were not indexed appropriately in PubMed, which complicated our initial search strategy. To carry out the search strategy, we have created and tested the blocks consecutively to gather the best results from each block, expanding and narrowing the search strategy. To assess the right direction of the search strategy, we have used fundamental literature, such as Choudhury & Asan 2020, Park et al. 2020 and Nagendran et al. 2020 as test sets, making sure the results of our search had common ground with these studies.

The resulting search string for Medline and PubMed in the individual blocks can be found in table 2 and describes the basis for other databases.

**Table 2:**
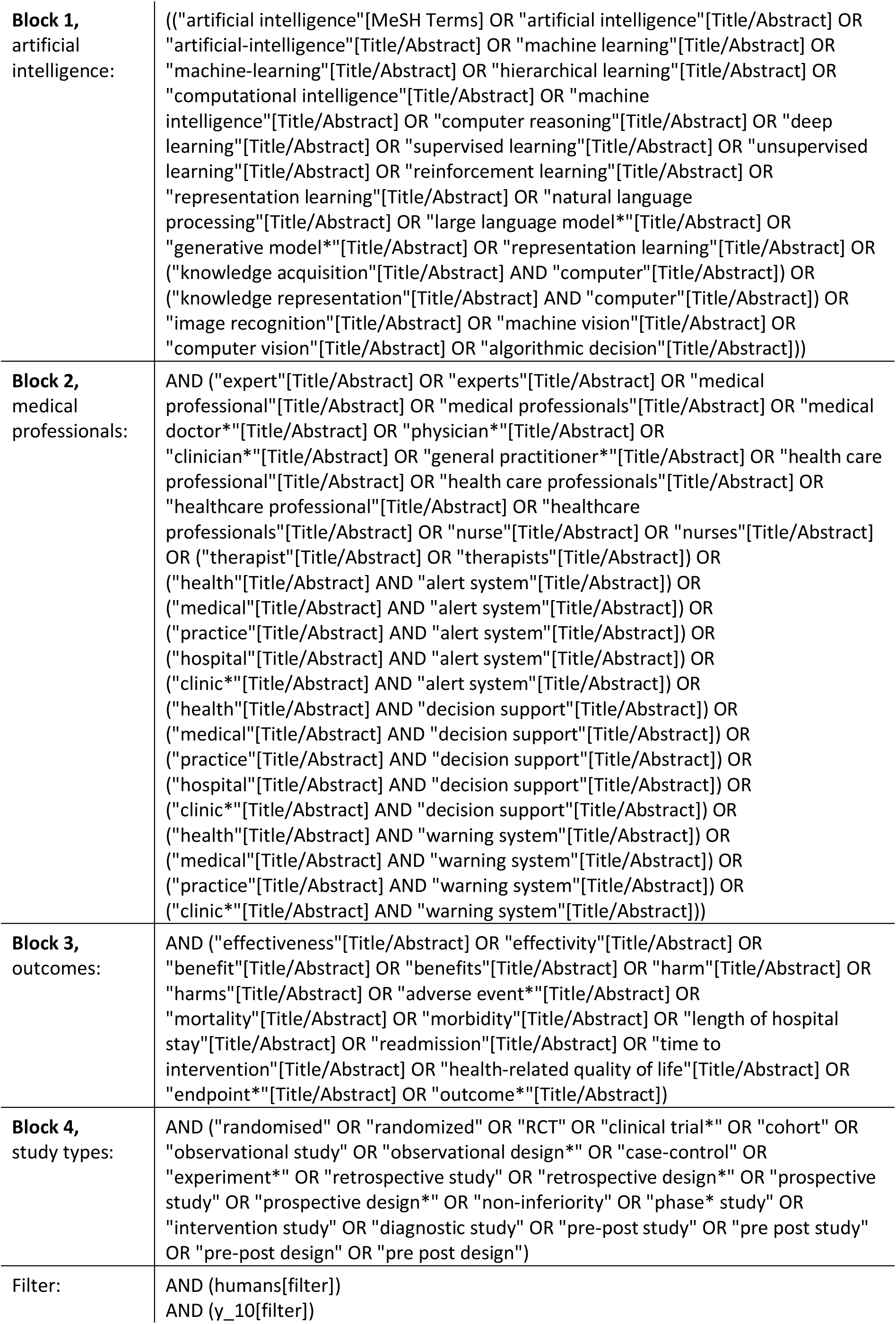
Search string blocks for Medline and PubMed (via PubMed)

### Types of studies to be included

For the systematic search peer reviewed interventional and observational studies published in German or English, 10 years retrospectively from the date of the search will be considered. For the search of grey literature, scientific reports published in German or English 10 years retrospectively from the date of the search will be considered. To extract potentially relevant studies from (systematic) reviews and meta-analyses, secondary studies will be gathered and screened. However, secondary studies will not be included in the synthesis.

In contrast to studies of effectiveness and safety, pure efficacy studies (e.g., focusing on algorithms accuracy) will be excluded as these outcomes are not directly relevant for patients. Patient-relevant outcomes will be defined according to the IQWiG method paper (2022). In addition, studies that used AI systems beyond our scopes, such as robotics (systems that support the implementation of decisions) will be excluded. Editorials, commentaries, letters, and other informal publication types will be excluded as well.

We will provide a list of all references screened in full text including exclusion reasons in the appendix of the final study.

### Participants

Our study is focusing on human patients without restriction of age or sex. Therefore, the input data for the algorithms must include real human data gathered either during routine care and saved for use in research or generated specifically for the individual study.

### Intervention

Out study is focusing on medical professionals utilizing an AI-related ADM system to address a clinical problem.

In our working definition, a medical professional is a qualified individual who has the authority to perform necessary medical procedures within their professional scope of practice. Their goal is to improve, maintain, or restore the health of individuals by examining, diagnosing, prognosticating, and/or treating clinical problems. This may include medical doctors, registered nurses, and other medical professionals. Clinical problems can encompass illnesses, injuries, and physical or mental disorders, among other conditions.

In our working definition, an AI-related ADM system is a clinical decision support system that either apply AI (e.g., relying on ML models) or that has been developed with the help of AI. Clinical decision support models without any involvement of AI will be excluded.

### Control

Medical professionals, as described in the working definition, are addressing a clinical problem without the support of an AI-related ADM system (standard care).

### Outcomes

Patient-relevant benefits and harms, according to the IQWiG method paper (2022), are gathered. These may include, for example, mortality, morbidity (with regard to complaints and complications), length of hospital stay, readmission, time to intervention and health-related quality of life.

### Study types

We will collect both interventional and observational studies, which may encompass randomised controlled trials, cohort studies, case-control studies, randomised surveys, retrospective and prospective studies, phase studies, as well as non-inferiority or diagnostic studies.

### Data extraction

Records arising from the literature search will be stored in the citation manager Citavi 6 (c) Swiss Academic Software. After removing duplicates, two reviewers will independently review all titles and abstracts via the browser application Rayyan (Ouzzani et al. 2016). Studies potentially meeting the inclusion criteria will then be screened in full text independently by two reviewers. Disagreements over eligibility of studies will be discussed and, if necessary, resolved by a third reviewer. Authors of the included studies will be contacted if clarification of their data or study methods is required. The PRISMA 2020 flow diagram (Page et al. 2021) will be used to keep the study selection process transparent.

Using a standardised data collection form, two reviewers will extract data independently from the included studies and will compare them for discrepancies. Missing data will be requested from study authors. Extracted data will include the following (Table 3).

**Table 3:**
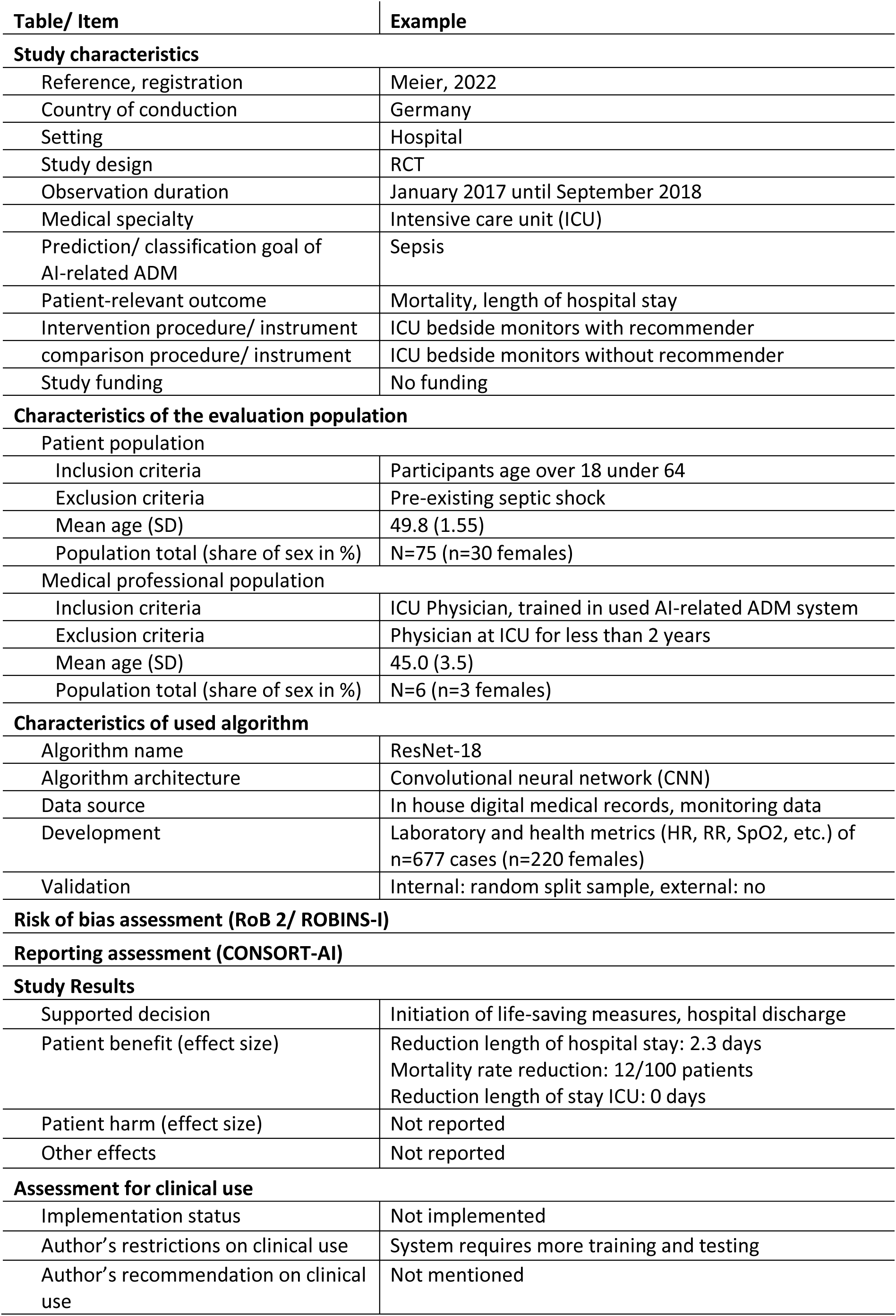
Study data to be extracted.

| Table/ Item | Example |
| --- | --- |
| **Study characteristics** |  |
| Reference, registration | Meier, 2022 |
| Country of conduction | Germany |
| Setting | Hospital |
| Study design | RCT |
| Observation duration | January 2017 until September 2018 |
| Medical specialty | Intensive care unit (ICU) |
| Prediction/ classification goal of AI-related ADM | Sepsis |
| Patient-relevant outcome | Mortality, length of hospital stay |
| Intervention procedure/ instrument | ICU bedside monitors with recommender |
| comparison procedure/ instrument | ICU bedside monitors without recommender |
| Study funding | No funding |
| **Characteristics of the evaluation population** |  |
| Patient population |  |
| Inclusion criteria | Participants age over 18 under 64 |
| Exclusion criteria | Pre-existing septic shock |
| Mean age (SD) | 49.8 (1.55) |
| Population total (share of sex in %) | N=75 (n=30 females) |
| Medical professional population |  |
| Inclusion criteria | ICU Physician, trained in used AI-related ADM system |
| Exclusion criteria | Physician at ICU for less than 2 years |
| Mean age (SD) | 45.0 (3.5) |
| Population total (share of sex in %) | N=6 (n=3 females) |
| **Characteristics of used algorithm** |  |
| Algorithm name | ResNet-18 |
| Algorithm architecture | Convolutional neural network (CNN) |
| Data source | In house digital medical records, monitoring data |
| Development | Laboratory and health metrics (HR, RR, SpO2, etc.) of n=677 cases (n=220 females) |
| Validation | Internal: random split sample, external: no |
| **Risk of bias assessment (RoB 2/ ROBINS-I)** |  |
| **Reporting assessment (CONSORT-AI)** |  |
| **Study Results** |  |
| Supported decision | Initiation of life-saving measures, hospital discharge |
| Patient benefit (effect size) | Reduction length of hospital stay: 2.3 days<br>Mortality rate reduction: 12/100 patients<br>Reduction length of stay ICU: 0 days |
| Patient harm (effect size) | Not reported |
| Other effects | Not reported |
| **Assessment for clinical use** |  |
| Implementation status | Not implemented |
| Author's restrictions on clinical use | System requires more training and testing |
| Author's recommendation on clinical use | Not mentioned |

### Risk of bias and quality assessment

Risk of bias will be assessed by using the revised Cochrane risk-of-bias tool for randomized trials (RoB 2) (Sterne et al. 2019) and the risk-of-bias in non-randomized studies for interventions (ROBINS-I) tool (Sterne et al. 2017). Disagreements between the authors over the risk of bias in the included studies will be resolved by discussion, or with involvement of a third author if necessary. Transparent reporting of the included studies will be assessed trough the Consolidated Standards of Reporting Trials interventions involving Artificial Intelligence (CONSORT-AI) extension by Liu et al. (2020). The CONSORT-AI extension includes 14 new items that were considered sufficiently important for AI interventions to be routinely reported in addition to the core CONSORT items by Schulz et al. (2010). CONSORT-AI aims to improve the transparency and completeness in reporting clinical trials for AI interventions. It will assist to understand, interpret, and critically appraise the quality of clinical trial design and risk of bias in the reported outcomes.

### Data synthesis

Given the expected likelihood of heterogeneity between studies in the different medical specialties in terms of outcome measures, study designs and interventions, we do not know if performing a meta-analysis will be possible. However, a systematic narrative synthesis will be provided of the results with an overview of the relevant effects for the outcomes, with information presented in the text and tables to summarise and explain the characteristics and findings of the included studies. The measures will be presented in absolute risks and risk differences. Studies of unclear or high risk of bias will not be excluded, as the authors assume that most potentially relevant studies will be of low methodological quality. This is also reported in recent reviews that examine the methodological quality of machine learning systems in the clinical setting (e.g., Nagendran et al. 2020). Nevertheless, the influence of the potential biases on the results will be discussed.

In addition to the synthesis trough Study 1, the results of this review will be discussed with experts (Study 2) and presented in the text.

### Study 2: Expert interviews

In order to meet the study objectives, and considering the findings of Study 1, we elicit experts’ experiences, knowledge, and their perceived benefits and risks of implementing AI-related ADM systems. By doing so, we hope to gain a practice-related understanding of the findings of Study 1 and identify barriers and needs regarding future research questions.

Based on the research questions and the findings of Study 1, the study and interview guide will be developed a priori. The study design and the interview guide will be submitted to the ethics committee of the University of Potsdam and pre-registered within the PROSPERO protocol for the systematic review. If necessary, post-registration changes to the protocol will be detailed and justified in the PROSPERO protocol. To ensure transparency of the qualitative research part of our review, we will follow the Standards for Reporting Qualitative Research (SRQR) (O’Brien et al. 2014).

### Study design

We will conduct semi-structured interviews. To analyse the interviews, we will use qualitative content analysis according to Mayring (2020). This systematic and rule-based approach will ensure that the analysis is understandable, comprehensible, and verifiable by others.

### Data collection

As our interview study aims to understand the practical implications of the findings of Study 1, we will use a purposive sampling strategy to select at least 3 interviewees according to their expertise through their function, profession, practice, and research in relation to the research questions (Mey & Mruck 2020: 322, Schreier 2020: 24). Therefore, we will combine experts of the following types: Auditing bodies responsible for auditing AI systems for safety (e.g., TÜV NORD IT Secure Communications GmbH & Co. KG), auditing bodies that support implementation procedures under scientific and regulatory conditions in the healthcare sector (e.g., VDI/VDE Innovation + Technik GmbH), and institutions that combine practical healthcare-related work and research with AI systems (e.g., Berlin Institute of Health at Charité Berlin). The experts will be recruited on the basis of publications, via professional networks, and by sending inquiries to the relevant organisations and institutions. Once relevant experts will have been identified, they will be contacted via e-mail or phone, informing about the specific research questions, and showing some exemplary findings from Study 1, to ask for participation and to set the scope of the interview. There is no reimbursement planned for the experts who participate in the interview.

Written informed consent will be obtained prior to each interview and participants will be fully informed in writing and verbally about the purpose, risks, and scope of the study.

Approximately one week prior to the interview, participants will be presented with the key findings of Study 1 and the interview guide, which they will be asked about in the interview. In addition to the questions on research interest, descriptive data will be collected on field of activity, specific expertise, organisation, length of involvement in the topic, as well as age and gender of the respective interviewee. Interviews should not exceed 45 minutes. All interviews will be audio recorded and automatically transcribed verbatim using Adobe Premiere Pro 2023 (version 23.1.0). The automated transcripts will be corrected manually by the interviewer.

Throughout the data collection process, the interviewer will keep a research diary to record initial impressions immediately after each interview and to encourage reflexivity about how their role in the researcher-participant relationship might influence the data collection process (Mayring 2020: 12).

### Data analysis and extraction

The transcripts and notes from the research diary will be transferred to the relevant memos of each transcript in MAXQDA Plus 2022 (version 22.2.0). All data analysis will be carried out by two independent researchers to ensure rigour and trustworthiness. In the event of disagreement between the two researchers on coding, we will revisit the data to develop a shared understanding of the consistent interpretations. The interviews will be analysed using qualitative content analysis according to Mayring (2020). The main categories will initially be developed deductively from the interview guide, which will be developed from the findings of Study 1. Additional major categories will be developed inductively from the data. The final code system, including illustrative interview quotes, will be part of the scientific publication. For this purpose, extracts from the transcript will be translated into English after coding, if necessary.

### Data management, data storage and data privacy

After being contacted by the interviewer via phone or e-mail, the experts will receive all relevant study documents via mail or e-mail. Verbal consent to receive the study documents will be obtained by phone or e-mail, and written consent will be obtained in the consent form before the interview takes place. All participants will be assured of confidentiality, anonymity of data, and the right to withdraw from the study or skip questions without negative consequences. Individual contact details will be stored on the interviewer’s password-protected computer until the time of the interview. Contact details will be deleted immediately after the interview. The audio recording is used only for quality assurance and will be deleted immediately after it is transcribed and compared. Transcripts will be pseudonymised by changing personal characteristics (such as names and addresses) and personal data, i.e., data that make indirect identification possible (e.g., places or institutions) to pseudonyms.

## Discussion

It is to be expected that there is a significant lack of suitable studies comparing healthcare professionals with and without AI-related ADM systems regarding patient-relevant outcomes. It is assumed that this is due to, first, the lack of approval regulations for AI systems, second, the prioritisation of technical and clinical parameters over patient-relevant outcomes in the development of study designs, and, third, the prioritisation of AI for supporting clinical processes (e.g., administration). In addition, it is to be expected that a large proportion of the studies to be identified are of rather poor methodological quality and provide results that are rather difficult to generalise. Although reporting guidelines such as the Consolidated Standards of Reporting Trials (CONSORT) Statement (Schulz et al. 2010) are well-known and widely used in medical and public health research, they do not necessarily correspond to the novel protocol and study designs that are relevant for the assessment of the research questions relevant here. The extension of the Reporting Guidelines for Clinical Study Reports of Interventions Using Artificial Intelligence (CONSORT-AI) (Liu et al. 2020) may fill the gap but this guideline is relatively new and not necessarily always applied.

Engaging in discussions with experts from AI safety audit organizations, healthcare implementation audit bodies, and institutions bridging healthcare practice and AI research, we will gain valuable insights into the implications of study 1’s findings (e.g., explanations for limited patient-relevant studies). Additionally, these discussions will illuminate the challenges and opportunities associated with the integration of AI-related ADM systems in healthcare practice and enlighten their benefits and harms for patients.

## Data Availability

All data produced in the present study are available upon reasonable request to the authors.

## List of abbreviations

ADM: Algorithmic Decision Making
AI: Artificial Intelligence
AUROC: Area Under the Receiver Operating Characteristic Curve
CENTRAL: Cochrane Central Register of Controlled Trials
CNN: Convolutional Neural Network
CONSORT: Consolidated Standards of Reporting Trials
CONSORT-AI: Consolidated Standards of Reporting Trials for Artificial Intelligence
CRD: Centre for Reviews and Dissemination
EMTREE: Embase subject headings
HR: Heart Rate
ICU: Intensive Care Unit
IEEE: Institute of Electrical and Electronics Engineers
IQWiG: German Institute for Quality and Efficiency in Healthcare
MECIR: Methodological Expectations of Cochrane Intervention Reviews
MeSH: Medical Subject Headings
ML: Machine Learning
nRCT: non Randomized Controlled Trial
PICO: Participants, Intervention, Control, Outcome
PRISMA: Preferred Reporting Items for Systematic Review and Meta-Analysis
PRISMA-P: Preferred Reporting Items for Systematic Review and Meta-Analysis Protocols
PROSPERO: International Prospective Register of Systematic Reviews
RCT: Randomized Controlled Trial
ResNet-18: A convolutional neural network that is 18 layers deep
RoB 2: Revised Cochrane Risk-of-Bias Tool for randomized trials
ROBINS-I: Risk-of-Bias in non-randomized studies for interventions
RR: Respiratory Rate
SD: Standard Deviation
SpO2: Oxygen Saturation
SRQR: Standards for Reporting Qualitative Research

## Declarations

## Ethics approval and consent to participate

Ethical approval for this study was obtained from the Ethics Committee of the University of Potsdam (103/2023).

## Consent for publication

Not applicable.

## Availability of data and materials

Not applicable.

## Acknowledgements

Not applicable

