## Supplement 1 for "Is artificial intelligence for medical professionals serving the patients? Protocol for a mixed method systematic review on patient-relevant benefits and harms of algorithmic decision-making"

### Search strings of databases

Database: MEDLINE and PubMed via PubMed, time of search: 27th of March 2024, Results: 1,923 hits

(("artificial intelligence"[MeSH Terms] OR "artificial intelligence"[Title/Abstract] OR "artificial-intelligence"[Title/Abstract] OR "machine learning"[Title/Abstract] OR "machine-learning"[Title/Abstract] OR "hierarchical learning"[Title/Abstract] OR "computational intelligence"[Title/Abstract] OR "machine intelligence"[Title/Abstract] OR "computer reasoning"[Title/Abstract] OR "deep learning"[Title/Abstract] OR "supervised learning"[Title/Abstract] OR "unsupervised learning"[Title/Abstract] OR "reinforcement learning"[Title/Abstract] OR "representation learning"[Title/Abstract] OR "natural language processing"[Title/Abstract] OR "large language model*"[Title/Abstract] OR "generative model*"[Title/Abstract] OR "representation learning"[Title/Abstract] OR ("knowledge acquisition"[Title/Abstract] AND "computer"[Title/Abstract]) OR ("knowledge representation"[Title/Abstract] AND "computer"[Title/Abstract]) OR "image recognition"[Title/Abstract] OR "machine vision"[Title/Abstract] OR "computer vision"[Title/Abstract] OR "algorithmic decision"[Title/Abstract]))

AND ("expert"[Title/Abstract] OR "experts"[Title/Abstract] OR "medical professional"[Title/Abstract] OR "medical professionals"[Title/Abstract] OR "medical doctor*"[Title/Abstract] OR "physician*"[Title/Abstract] OR "clinician*"[Title/Abstract] OR "general practitioner*"[Title/Abstract] OR "health care professional"[Title/Abstract] OR "health care professionals"[Title/Abstract] OR "healthcare professional"[Title/Abstract] OR "healthcare professionals"[Title/Abstract] OR "nurse"[Title/Abstract] OR "nurses"[Title/Abstract] OR ("therapist"[Title/Abstract] OR "therapists"[Title/Abstract]) OR ("health"[Title/Abstract] AND "alert system"[Title/Abstract]) OR ("medical"[Title/Abstract] AND "alert system"[Title/Abstract]) OR ("practice"[Title/Abstract] AND "alert system"[Title/Abstract]) OR ("hospital"[Title/Abstract] AND "alert system"[Title/Abstract]) OR ("clinic*"[Title/Abstract] AND "alert system"[Title/Abstract]) OR ("health"[Title/Abstract] AND "decision support"[Title/Abstract]) OR ("medical"[Title/Abstract] AND "decision support"[Title/Abstract]) OR ("practice"[Title/Abstract] AND "decision support"[Title/Abstract]) OR ("hospital"[Title/Abstract] AND "decision support"[Title/Abstract]) OR ("clinic*"[Title/Abstract] AND "decision support"[Title/Abstract]) OR ("health"[Title/Abstract] AND "warning system"[Title/Abstract]) OR ("medical"[Title/Abstract] AND "warning system"[Title/Abstract]) OR ("practice"[Title/Abstract] AND "warning system"[Title/Abstract]) OR ("clinic*"[Title/Abstract] AND "warning system"[Title/Abstract]))

AND ("effectiveness"[Title/Abstract] OR "effectivity"[Title/Abstract] OR "benefit"[Title/Abstract] OR "benefits"[Title/Abstract] OR "harm"[Title/Abstract] OR "harms"[Title/Abstract] OR "adverse event*"[Title/Abstract] OR "mortality"[Title/Abstract] OR "morbidity"[Title/Abstract] OR "length of hospital stay"[Title/Abstract] OR "readmission"[Title/Abstract] OR "time to intervention"[Title/Abstract] OR "health-related quality of life"[Title/Abstract] OR "endpoint*"[Title/Abstract] OR "outcome*"[Title/Abstract])

AND ("randomised" OR "randomized" OR "RCT" OR "clinical trial*" OR "cohort" OR "observational study" OR "observational design*" OR "case-control" OR "experiment*" OR "retrospective study" OR "retrospective design*" OR "prospective study" OR "prospective design*" OR "non-inferiority" OR "phase* study" OR "intervention study" OR "diagnostic study" OR "pre-post study" OR "pre post study" OR "pre-post design" OR "pre post design")

AND (y_10[Filter]) AND (humans[Filter])

Database: Embase via Elsevier, time of search: 27th of March 2024, Results: 1,000 hits

('artificial intelligence'/exp OR 'artificial intelligence':ti,ab OR 'artificial-intelligence':ti,ab OR 'machine learning':ti,ab OR 'machine-learning':ti,ab OR 'hierarchical learning':ti,ab OR 'computational intelligence':ti,ab OR 'machine intelligence':ti,ab OR 'computer reasoning':ti,ab OR 'deep learning':ti,ab OR 'supervised learning':ti,ab OR 'unsupervised learning':ti,ab OR 'reinforcement learning':ti,ab OR 'natural language processing':ti,ab OR 'large language model*':ti,ab OR 'generative model*':ti,ab OR 'representation learning':ti,ab OR ('knowledge acquisition':ti,ab AND 'computer':ti,ab) OR ('knowledge representation':ti,ab AND 'computer':ti,ab) OR 'image recognition':ti,ab OR 'machine vision':ti,ab OR 'computer vision':ti,ab OR 'algorithmic decision':ti,ab)

AND ('expert':ti,ab OR 'experts':ti,ab OR 'medical professional':ti,ab OR 'medical professionals':ti,ab OR 'medical doctor*':ti,ab OR 'physician*':ti,ab OR 'clinician*':ti,ab OR 'general practitioner*':ti,ab OR 'health care professional':ti,ab OR 'health care professionals':ti,ab OR 'healthcare professional':ti,ab OR 'healthcare professionals':ti,ab OR 'nurse':ti,ab OR 'nurses':ti,ab OR 'therapist':ti,ab OR 'therapists':ti,ab OR ('health':ti,ab AND 'alert system':ti,ab) OR ('medical':ti,ab AND 'alert system':ti,ab) OR ('practice':ti,ab AND 'alert system':ti,ab) OR ('hospital':ti,ab AND 'alert system':ti,ab) OR ('clinic*':ti,ab AND 'alert system':ti,ab) OR ('health':ti,ab AND 'decision support':ti,ab) OR ('medical':ti,ab AND 'decision support':ti,ab) OR ('practice':ti,ab AND 'decision support':ti,ab) OR ('hospital':ti,ab AND 'decision support':ti,ab) OR ('clinic*':ti,ab AND 'decision support':ti,ab) OR ('health':ti,ab AND 'warning system':ti,ab) OR ('medical':ti,ab AND 'warning system':ti,ab) OR ('practice':ti,ab AND 'warning system':ti,ab) OR ('clinic*':ti,ab AND 'warning system':ti,ab))

AND ('effectiveness':ti,ab OR 'effectivity':ti,ab OR 'benefit':ti,ab OR 'benefits':ti,ab OR 'harm':ti,ab OR 'harms':ti,ab OR 'adverse event*':ti,ab OR 'mortality':ti,ab OR 'morbidity':ti,ab OR 'length of hospital stay':ti,ab OR 'readmission':ti,ab OR 'time to intervention':ti,ab OR 'health-related quality of life':ti,ab OR 'endpoint*':ti,ab OR 'outcome*':ti,ab)

AND ('randomised':ti,ab OR 'randomized':ti,ab OR 'rct':ti,ab OR 'clinical trial*':ti,ab OR 'cohort':ti,ab OR 'observational study':ti,ab OR 'observational design*':ti,ab OR 'case-control':ti,ab OR 'experiment*':ti,ab OR 'retrospective study':ti,ab OR 'retrospective design*':ti,ab OR 'prospective study':ti,ab OR 'prospective design*':ti,ab OR 'non-inferiority':ti,ab OR 'phase* study':ti,ab OR 'intervention study':ti,ab OR 'diagnostic study':ti,ab OR 'pre post study':ti,ab OR 'pre-post study':ti,ab OR 'pre post design':ti,ab OR 'pre-post design':ti,ab)

AND [humans]/lim AND [clinical study]/lim AND [embase]/lim AND [2014-2024]/py AND [article]/lim

Database: IEEE Xplore via ieeexplore.ieee.org, time of search: 27th of March 2024, Results: 77 hits

((("artificial intelligence" OR "artificial-intelligence" OR "machine learning" OR "machine-learning" OR "algorithmic decision" OR "hierarchical learning" OR "computational intelligence" OR "machine intelligence" OR "computer reasoning" OR "deep learning" OR "supervised learning" OR "unsupervised learning" OR "reinforcement learning" OR "representation learning" OR "natural language processing" OR "large language model" OR "large language models" OR "generative models" OR "representation learning" OR "image recognition" OR "machine vision" OR "computer vision")

AND ("length of hospital stay" OR "readmission" OR "time to intervention" OR "health-related quality of life" OR "endpoint" OR "endpoints" OR "outcome" OR "outcomes")

AND ("randomised" OR "randomized" OR "RCT" OR "clinical trial" OR "clinical trials" OR "cohort" OR "observational study" OR "observational design" OR "observational designs" OR "case-control" OR "experiment" OR "experiments" OR "randomised survey" OR "randomized survey" OR "retrospective study" OR "retrospective design" OR "retrospective designs" OR "prospective study" OR "prospective design" OR "prospective designs" OR "non-inferiority" OR "phase study" OR "phases study" OR "intervention study" OR "diagnostic study" OR "pre-post study" OR "pre post study" OR "pre-post design" OR "pre post design")

AND ("expert" OR "experts" OR "medical professional" OR "medical professionals" OR "medical doctor" OR "medical doctors" OR "physician" OR "physicians" OR "clinician" OR "clinicians" OR "general practitioner" OR "health care professional" OR "health care professionals" OR "healthcare professional" OR "healthcare professionals" OR "nurse" OR "nurses" OR "therapist" OR "therapists" OR ("health" AND "warning system") OR ("clinical" AND "decision support") OR ("health" AND "alert system") OR ("medical" AND "alert system") OR ("practice" AND "alert system") OR ("hospital" AND "alert system") OR ("clinical" AND "alert system") OR ("health" AND "decision support") OR ("medical" AND "decision support") OR ("practice" AND "decision support") OR ("hospital" AND "decision support") OR ("medical" AND "warning system") OR ("practice" AND "warning system") OR ("clinical" AND "warning system"))

AND ("effectiveness" OR "effectivity" OR "benefit" OR "benefits" OR "harm" OR "harms" OR "adverse event" OR "adverse events" OR "mortality" OR "morbidity" OR "length of hospital stay" OR "readmission" OR "time to intervention" OR "health-related quality of life" OR "endpoint" OR "endpoints" OR "outcome" OR "outcomes")))

Filters Applied: Journals, Early Access Articles, Magazines, Publication Date 2014 – 2024

Database: ICTRP and clinicaltrails.gov via CENTRAL - Cochrane Central Register of Controlled Trials, time of search: 27th of March 2024, Results: 166 hits

MeSH descriptor: [Artificial Intelligence] explode all trees OR ("artificial intelligence" OR "artificial-intelligence" OR "machine learning" OR "machine-learning" OR "hierarchical learning" OR "computational intelligence" OR "machine intelligence" OR "computer reasoning" OR "deep learning" OR "supervised learning" OR "unsupervised learning" OR "reinforcement learning" OR "representation learning" OR "natural language processing" OR "large language model" OR "large language models" OR "generative model" OR "generative models" OR "representation learning" OR "image recognition" OR "machine vision" OR "computer vision" OR "algorithmic decision"):ti,ab,kw OR ((knowledge acquisition AND computer) OR (knowledge representation AND computer)):ti,ab,kw

AND "expert" OR "experts" OR "medical professional" OR "medical professionals" OR "medical doctor" OR "medical doctors" OR "physician" OR "physicians" OR "clinician" OR "clinicians" OR "general practitioner" OR "health care professional" OR "health care professionals" OR "healthcare professional" OR "healthcare professionals" OR "nurse" OR "nurses" OR "therapist" OR "therapists" OR ("health" AND "warning system") OR ("clinical" AND "decision support") OR ("health" AND "alert system") OR ("medical" AND "alert system") OR ("practice" AND "alert system") OR ("hospital" AND "alert system") OR ("clinical" AND "alert system") OR ("health" AND "decision support") OR ("medical" AND "decision support") OR ("practice" AND "decision support") OR ("hospital" AND "decision support") OR ("medical" AND "warning system") OR ("practice" AND "warning system") OR ("clinical" AND "warning system"):ti,ab,kw

AND ("effectiveness" OR "effectivity" OR "benefit" OR "benefits" OR "harm" OR "harms" OR "adverse event" OR "adverse events" OR "mortality" OR "morbidity" OR "length of hospital stay" OR "readmission" OR "time to intervention" OR "health-related quality of life" OR "endpoint" OR "endpoints" OR "outcome" OR "outcomes"):ti,ab,kw

AND ("randomised OR "randomized" OR "RCT" OR "clinical trial" OR "clinical trials" OR "clinical trials" OR "cohort" OR "observational study" OR "observational design" OR "observational designs" OR "case-control" OR "experiment" OR "experiments" OR "retrospective study" OR "retrospective design" OR "retrospective designs" OR "prospective study" OR "prospective design" OR "prospective designs" OR "non-inferiority" OR "phase study" OR "phases study" OR "intervention study" OR "diagnostic study" OR "pre-post study" OR "pre post study" OR "pre-post design" OR "pre post design"):ti,ab,kw
